# Divide in Vaccine Belief in COVID-19 Conversations: Implications for Immunization Plans

**DOI:** 10.1101/2020.07.23.20160887

**Authors:** Aman Tyagi, Kathleen M. Carley

## Abstract

The development of a viable COVID-19 vaccine is a work in progress, but the success of the immunization campaign will depend upon public acceptance. In this paper, we classify Twitter users in COVID-19 discussion into vaccine refusers (anti-vaxxers) and vaccine adherers (vaxxers) communities. We study the divide between anti-vaxxers and vaxxers in the context of whom they follow. More specifically, we look at followership of 1) the U.S. Congress members, 2) four major religions (Christianity, Hinduism, Judaism and Islam), 3) accounts related to the healthcare community, and 4) news media accounts. Our results indicate that there is a partisan divide between vaxxers and anti-vaxxers. We find a religious community with a higher than expected fraction of anti-vaxxers. Further, we find that the variance of vaccine belief within the news media accounts operated by Russian and Iranian governments is higher compared to news media accounts operated by other governments. Finally, we provide messaging and policy implications to inform the COVID-19 vaccine and future vaccination plans.

## 1 Introduction

Social media platforms such as Twitter have become an important medium for organizing around complex socio-political issues. Recently, one such issue is the discussion about COVID-19. There have been many different studies conducted about COVID-19 in social media, particularly surrounding disinformation [1, 15, 23]. In this paper, we focus on classifying Twitter users in COVID-19 discussion into vaccine refuser (anti-vaxxers) and vaccine adherer (vaxxers) communities. As scientists and researchers rush to invent a vaccine to contain the virus, people opposing vaccination could present a significant challenge. An understanding of the anti-vaxxers and whom they follow would help deliver targeted messaging and help build policy tools to develop confidence in vaccination.

Recent works have found that there is a partisan divide among vaxxers and anti-vaxxers. In a recent study, Walter et. al [27] suggested that there is partisan polarization in vaccine discourse. A study by Broniatowski and colleagues [9] concluded that Russian bots (automated accounts) and trolls amplified the vaccine debate creating confusion about vaccines. Moreover, [12] concluded that during the US presidential elections vaccine opponents are significantly more likely to mention President Trump.

In this study, instead of solely focusing on the partisan divide between vaxxers and anti-vaxxers, we also look at other characteristics that could explain vaccine beliefs. First, we further explore partisan polarization based on the type of news and U.S. Congress members people follow. Second, we explore cultural differences by examining the followership of different religions, and the news media accounts owned or operated by different countries around the world. Finally, we look at the followership of doctors and medical institutions to better understand how healthcare community members could help in delivering more harmonious vaccine messaging.

We present how following different religious institutions, doctors, medical associations and institutes, and news media could predict vaccine belief, specifically in the context of the COVID-19 debate. We primarily investigate the following question: *How an individual’s vaccine belief differs with respect to influencers that individual follows?* To this end, we look at Twitter users in COVID-19 debate that mention vaccine-related words and classify them into vaxxers and anti-vaxxers using a state-of-the-art method described in §2.2. We then collect followers of Twitter influencers from four different groups:

1. *Political Inclination*: US Democrat and Republican Congress members
2. *Religious Inclination*: Christian, Islam, Judaism, and Hinduism
3. *Healthcare*: doctors, medical organizations and institutes
4. *News Media*: news media sources owned fully or partially by different countries (state media) and right leaning, left leaning, and mixed audience news media as defined by Pew Research Centre’s study [17].

We find the fraction of Anti-Vaxxers and Vaxxers that follow the above groups. We also train logistic regression models to understand the effect on each individual’s vaccine belief by following different entities. This helps us understand how the divide within each group can explain vaccine beliefs.

We describe our dataset including manual collection for each group in §2.1. Next, we discuss our stance classification method in §2.2. Our results (§3) suggest that an individual following more republicans than democrats is more likely to be an anti-vaxxer. This is a better predictor than the followership pattern of religious, healthcare, or news media groups. Through this research, we 1) provide further evidence that how vaccination has become a partisan issue, 2) find direct influencers in anti-vaxxers debate, and 3) provide public health and policy implications for future COVID-19 vaccine.

## 2 Methods

To answer our research questions we use multiple data sources and models. Figure 1 describes the overview of our data collection and methods. We begin by describing our COVID-19 related tweets and keyword filtration to get vaccine-related discourse in §2.1. Next, we describe our stance detection method to classify users into anti-vaxxers and vaxxers in §2.2. Lastly, we present our logistic regression models in §2.3.

**Fig. 1.**
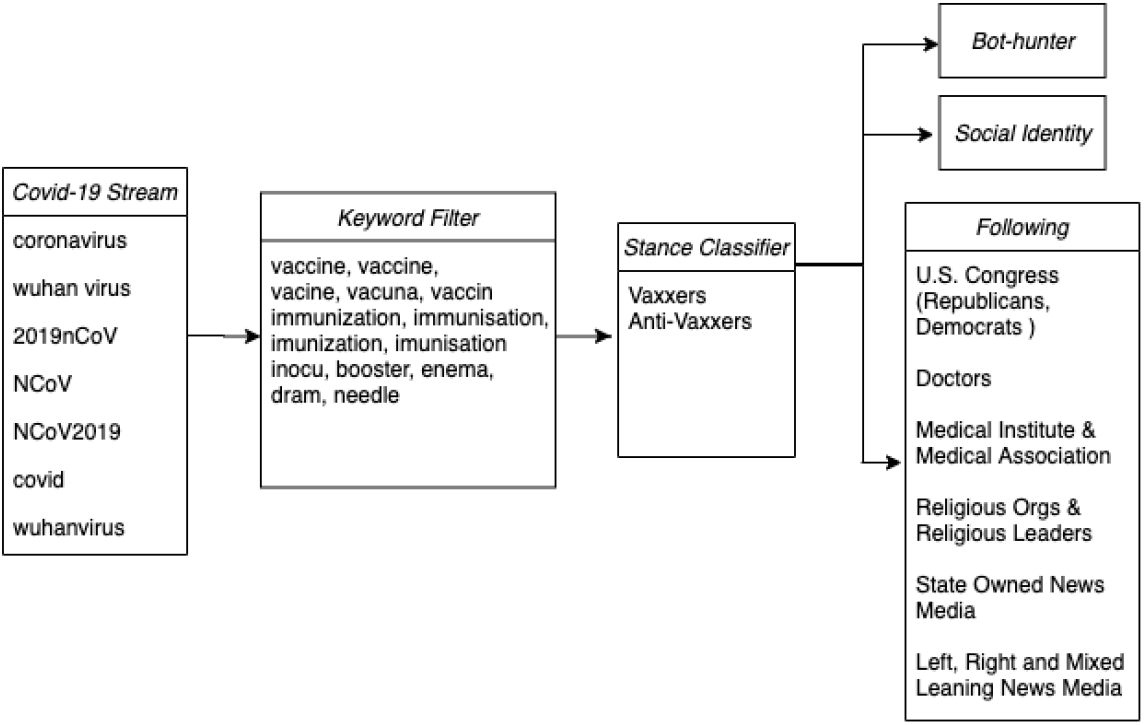
Data collection and methods pipeline. From left to right: COVID-19 Twitter stream with all collection keywords, keyword filter for vaccine related tweets, stance classifier, Bot-hunter [3, 4], social identity [6] and account groups used in our analysis.

### 2.1 Data Collection

We collected realtime tweets using Twitter’s standard API^1^ with COVID-19 related keywords ^2^. Our dataset was collected between February 1st, 2020 to April 30th, 2020. We then de-duplicated the collected tweets to remove any tweets collected more than once during everyday collection. Overall, we collected 227M unique tweets and retweets from 27M unique users. In this paper, we are primarily concerned with users tweeting in English and we do recognize that our selected keywords bias our collection.

Next, we filtered the tweets containing vaccine-related synonyms or their common mis-spellings. After the filtration, our Twitter dataset contained 3.9M tweets from 1.9M unique users. We then used a stance detection algorithm to find anti-vaxxers and vaxxers.

### 2.2 Stance Detection

Labeling each user as a vaxxer or an anti-vaxxer is a non-trivial task. The broader field of labeling users based on the position the user takes on a particular topic is called *stance mining* [22]. We use state-of-the-art stance mining method which uses weak supervision to find anti-vaxxers and vaxxers [19]. The model uses text signals from Tweets along with retweet and hashtag network features using a co-training approach with label propagation [28] and text classification. A set of seed hashtags are provided as a pro and anti stance signals to the model. The model then labels seed users based on the usage of these seed hashtags at the end of the tweet (endtags). The labeled and unlabeled users are then taken as input to the co-training algorithm. In each step, a combined user-retweet and user-hashtag network is used to propagate labels to unlabelled users. Concurrently, the text classifier uses the seed user’s tweets to train an SVM [10] based text classifier to predict unlabeled users. A common set from text classification and label propagation of highly confident labels are then used as seed labels for the next iteration. The final classification is based on the prediction of the joint model using the combined confidence scores.^3^ The model has been shown to be above 80% accurate with multiple datasets.

We select hashtag *VaccineInjury* and *SIDS* as anti-vaxxer seed hastags and *#VaccinateYourKids* and *#VaccineWorks* as vaxxers seed hashtags. Hashtags *VaccineInjury* and *SIDS* has been shown to be used mostly by anti-vaxxers [12]. We found similar results on using other anti-vaxxers hashtags reported in [12]. We use *VaccinateYourKids* and *VaccineWorks* as vaxxers hashtags because of their semantics. Out of the 1.92M users talking about vaccine in context of COVID-19, we classified 490k as anti-vaxxers and 1.43M as vaxxers. In Supplementary 1 we discuss the results from manual evaluation of a large sample. We observe that the average precision for the large sample is 82.6%.

### 2.3 Regression Model

We use a logistic regression model to study the effect of following different groups^4^ We then map the followers of different entities within these groups to check which entities are a better predictor of vaccine beliefs.

Logistic regression models typically require little or no multicollinearity. To overcome multicollinearity, we define a model for each of the four categories defined in §1. In figure 4 we report the correlation matrix for each of the independent variables that we define below. In our regression models, the number of observations is very large (*N* = 1849981) and hence we do not consider the p-values. With very large number of observations p-values tend towards zero [20] and hence we rely on regression coefficients and standard errors.

Previous research has concluded that discussion on different socio-economic topics has been amplified by automated accounts [9, 5, 26]. To control for bot-like accounts in our analysis, we used CMU’s Bot-Hunter [3, 4]. Bot-hunter’s output is a probability measure of bot-like behavior of an account. In each of our models described below, we use this probability output as 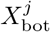 associated with account *j*. Also, we define 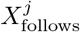 as the number of accounts followed by the user *j* at the time of first occurrence of the user in vaccine filtered tweets. We use the labels from our stance detection output as the independent variable, we define *l*^*j*^ as log odds of user *j* labeled as vaxxer.

Furthermore, a Twitter account could be a news media account *e*.*g*. a press reporter or a news agency, or an organizational entity such as the government, or a celebrity. As these types of accounts could potentially bias our regression model, we remove these accounts using a pre-trained model from [6]. The classification model in [6] was trained on the users’ tweets and personal descriptions using LSTM network [13] with attention mechanism [2]. The test accuracy reported in the study for the held-out data set is 91.6%. The model classifies each account into reporter, celebrity, news agency, company, government, sports-related and normal person’s account. The model was able to classify 96% of the accounts into normal accounts. In our regression model, we would only use these normal accounts to study the difference in beliefs. We define other dependent variables and the equation for each group below.

#### Political Inclination

We collected a list of official Twitter handles of all senators and representatives. For each vaxxer or an anti-vaxxer we find the number of republicans and democrats followed by the user ^5^. Equation 1 shows our reqression formula. We define, 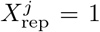 if the user follows strictly more number of Republican parliamentarians than Democrat parliamentarians otherwise 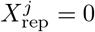. Similarly, 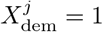 if the user follows strictly more number of Democrat parliamentarians than Republican parliamentarians otherwise 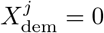.

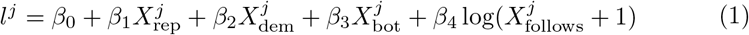

#### Religious Inclination

To compare the effect of following different religious groups on vaccine belief, we observe how vaxxers and anti-vaxxers follow religious accounts. We limit our analysis to four major religions, namely 1) Christianity, 2) Hinduism, 3) Judaism and, 4)Islam. We manually search for religious leaders and institutions on Twitter using keywords associated with each of these religions^6^. We then map the followers of each of these accounts with the accounts we labeled as vaxxer or anti-vaxxer. In Equation 2, we define 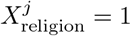, for each religion, if the user *j* follows strictly more accounts from that religion compared to other religions otherwise 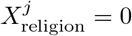

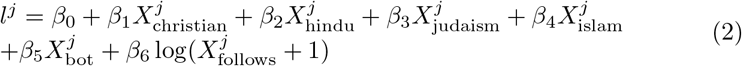

#### Healthcare

Similar to our religious inclination analysis, we observe how vaxxers and anti-vaxxers follow healthcare-related accounts. We manually searched for doctors, medical institutes, and medical associations on Twitter and Google. In Equation 3, we define 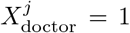, if the user *j* follows strictly more doctors than medical institutes and associations otherwise 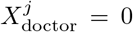. Similarly, we define 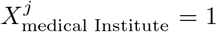 if the user *j* follows strictly more medical institutes and associations than doctors otherwise 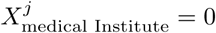.

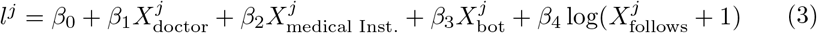

#### News Media

Finally, we compare the effect of following different news media types as defined by pew research [17]. Pew Research’s study combined a list of 30 most popular news media accounts. The study divides these news media sources into left-leaning, right-leaning, or mixed based on whether people supporting democrats, republicans, or both trust the sources respectively. We observe how vaxxers and anti-vaxxers are following Twitter handles of these 30 accounts ^7^. We then mapped the followers of each of these accounts with the accounts we labeled as vaxxer or anti-vaxxer. In Equation 4, we define 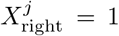, if the user *j* follows strictly more right-leaning accounts compared to other left and mixed audience accounts, 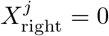 otherwise. Similarly, we define 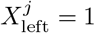 and 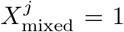, if the user *j* follows strictly more left-leaning and mixed accounts respectively and equal to 0 otherwise.

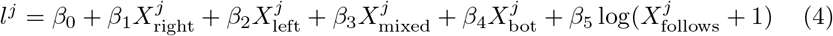

In news media category, we also define state media accounts as news sources that are partially owned or operated by a country’s government. We collected a list of 105 state media accounts from eight major countries. We then analyzed how many anti-vaxxers and vaxxers are following these accounts.

## 3 Results

We first examine the followers of each group for the fraction of vaxxers and anti-vaxxers and then describe our regression results. In Table 1 we report the percentage of vaxxers and anti-vaxxers and then describe our regression results. In Table 1 we report the percentage of anti-vaxxers and vaxxers following at least one account in each group. As compared to other groups, the highest fraction anti-vaxxers follow at least one Republican congress member. On the other hand, the highest fraction of vaxxers follow at least one left-leaning news media. Right-leaning news media and Republican congressperson show the highest difference between the percentage of anti-vaxxers and vaxxers. Similarly, Hindu religion-related and state media accounts show the lowest decrease in the percentage of vaxxers and anti-vaxxers. Moreover, vaxxers or anti-vaxxers are more likely to follow at least one Medical institute or association when compared to individual doctors. To examine the beliefs of the followers, we define the metric *η* in Equation 5. We then look into each of the four groups defined in §1 and how *η* changes for entities within these groups. For example, we would look into the *η* value for each of the Republican Congress member, and then compare the mean and standard deviation value of *η* for Republican members with Democrat members. If for a group −1 *<*= *η <* 0 then more vaxxers follow that group and if 0 *< η <*= 1 then more anti-vaxxers follow the group. To compare which users/groups have more vaxxers or anti-vaxxers compared to the expected value, we define *η*_*exp*_ value using the entire dataset with 490k anti-vaxxers and 1.43M vaxxers which comes out to be −0.488.

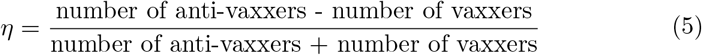

**Table 1.** Percentage of anti-vaxxers and vaxxers following at least one account from different groups and the difference between the percentage of anti-vaxxers and vaxxers. Number of accounts in each group is in parenthesis.

|  | % of Anti-Vaxxers | % of Vaxxers | Difference |
| --- | --- | --- | --- |
| Hindu (150) | 5.23 | 10.1 | -4.87 |
| Judaism (150) | 7.04 | 4.58 | 2.46 |
| Christian (150) | 17.4 | 6.30 | 11.1 |
| Islam (150) | 5.99 | 6.64 | -0.65 |
| Democrat (282) | 19.2 | 15.9 | 3.30 |
| Republican (249) | **32.2** | 9.41 | **22.8** |
| State Media (106) | 15.3 | 24.6 | -9.30 |
| Medical Assc. & Inst. (50) | 15.7 | 17.0 | -1.30 |
| Doctors (50) | 6.81 | 6.40 | 0.41 |
| Left Leaning Media (17) | 25.6 | **27.9** | -2.30 |
| Mixed Audience Media (7) | 15.2 | 15.0 | 0.20 |
| Right Leaning Media (6) | 28.5 | 6.26 | 22.2 |

### Political Inclination

Figure 2 visualizes the average *η* value of Republican and Democrat Congress members for each state in the senate and the house of representatives. In both the houses, we see a clear difference between Republican and Democrat. There are more anti-vaxxers following Republicans than Democrats. Interestingly, this pattern remains consistent for senators of all parts of the US except for Florida, Pennsylvania, and Maine. However, in the case of representatives, only among the followers of Nevada and New Jersey representatives from the Republican party have more Vaxxers than Anti-Vaxxers. For most of the other states, there is no abrupt difference between the two houses. States such as the Dakotas, Louisiana, Wyoming, and Arizona have a very high fraction of Anti-vaxxers among the followers of the Republican party leaders.

**Fig. 2.**
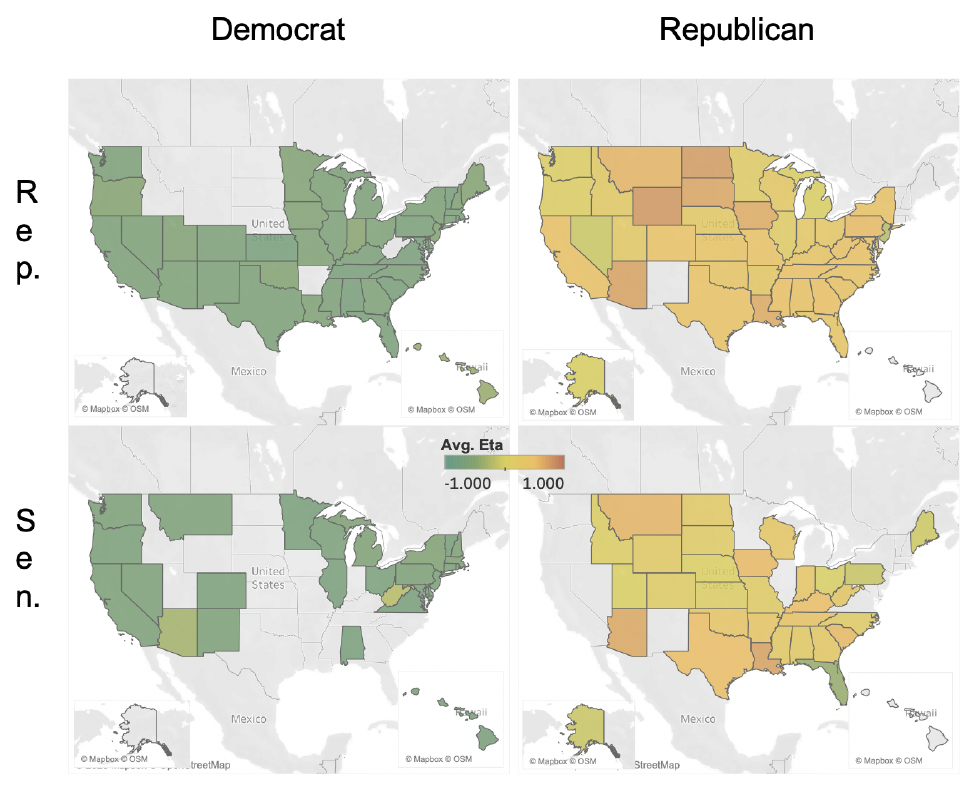
Average *η* value for Republican and Democrat members of Senate and House of Representatives for 116th U.S. Congress. The color scale (middle) represent *η* = −1 (green), *η* = 0 (yellow) and *η* = 1 (red). Lack of color represents no member of that respective party is in U.S. Congress. Rep. Duncan Hunter, Rep. Chris Collins, Rep. Lance Gooden, and Rep. Sean Duffy from the Republican party are not part of this analysis due to the absence of an active official Twitter account.

### Religion

The quantiles for *η* values for accounts related to four major religions is presented in Figure 3 (top-left). Our results suggest that accounts related to Hinduism have the least number of anti-vaxxers followed by Islam. Although we have a very large standard deviation in *η* values for Christianity, the median *η* value for Christian related accounts is still much higher than the *η*_*exp*_. This indicates that some Christianity related accounts are more popular among anti-vaxxers. For accounts related to Judaism, we have a very large standard deviation in *η* values with the median *η* a bit lower than *η*_*exp*_.

**Fig. 3.**
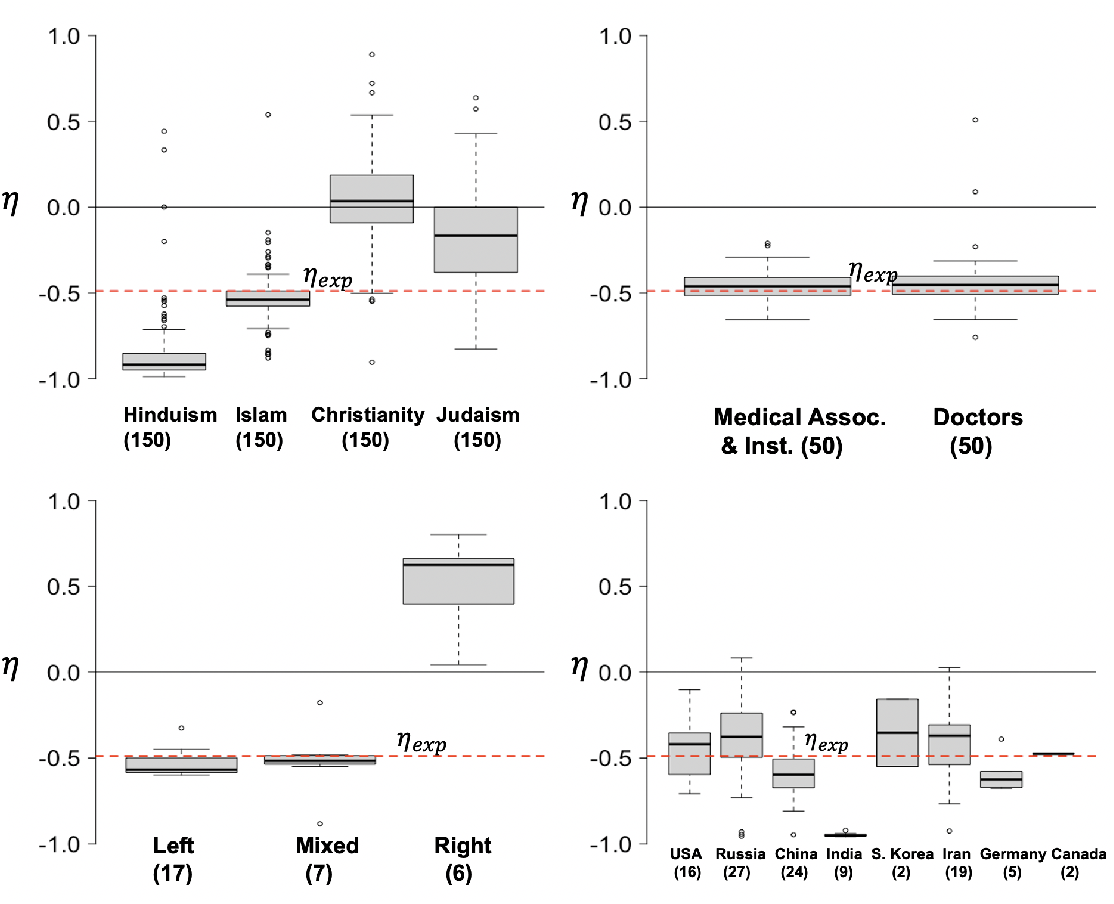
Boxplot for *η* valuea for news media as labeled by the type of audience by Pew Research [17] (bottom-left), news media partially or fully owned by countries (bottom-right), healthcare professionals and associations (top-right), and different religions (top-left). The number of accounts in each group is in parenthesis.

**Fig. 4.**
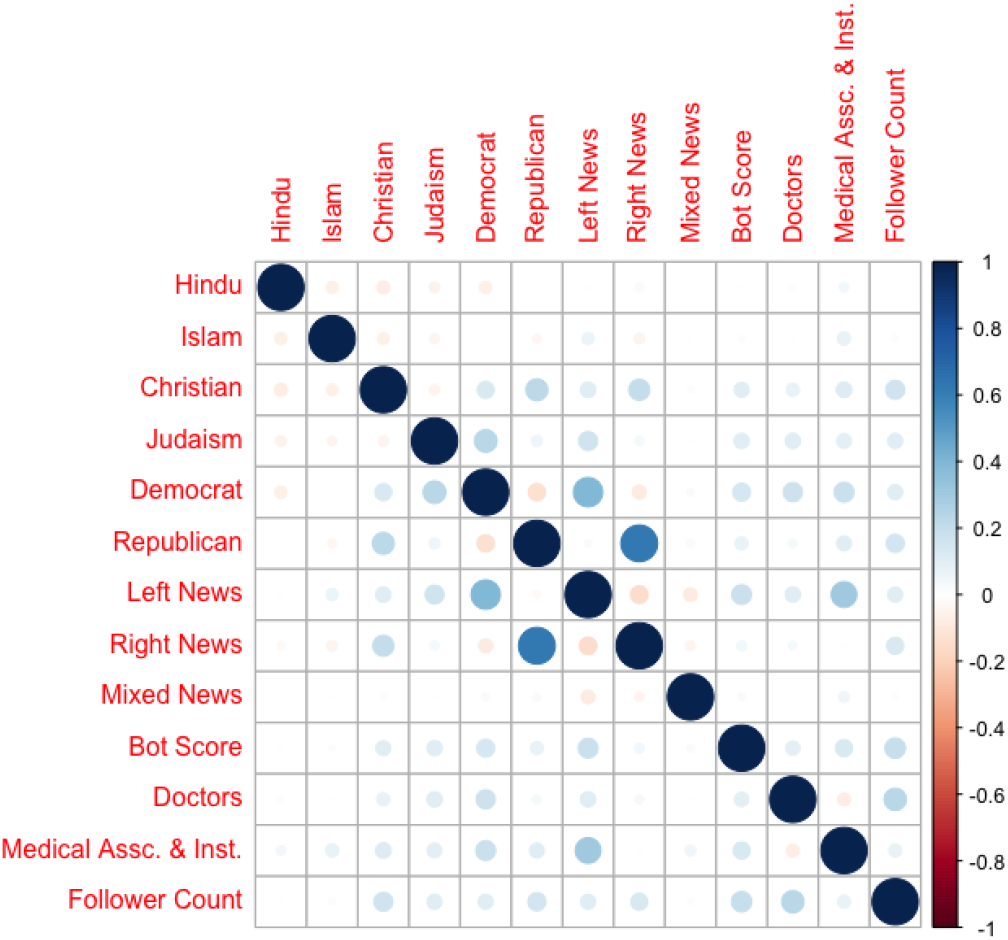
Correlation matrix of variables defined in §2.3

### Healthcare

For both doctors and medical association and institutes, the median *η* value is closer to *η*_*exp*_. The standard deviation for doctors is higher than compared to the other group, indicating that most doctors are more popular among vaxxers, while few are have more anti-vaxxers compared to vaxxers.

### News Media

In Figure 3 (bottom-left), we see a clear difference in median *η* value for right-leaning news media as compared to left or mixed audience media. This finding is consistent with the results we found about political inclination. Next, we group all the state media accounts by country and present the median *η* values in Figure 3 (bottom-right). Indian state media accounts stand out with a very high concentration of vaxxers. Although in this study we have a higher number of Twitter accounts operated by Russian and Iranian government, the standard deviation for both these countries is much higher than for other countries. Indicating that there exists a divide in how many vaxxers and anti-vaxxers are following the news media accounts owned or operated by a particular country.

### Regression Results

In Table 2 we present the mean area under the receiver operating characteristics of the models discussed in §2.3. Although the number of independent variables is higher than the religious inclination model, the political inclination model has higher accuracy than other individual models which is closely followed by the news media model. This suggests that the divide in vaccine belief is strongly predicted by the political divide as compared to the religious divide. In Supplementary 1, we present the estimates and the standard errors of each model (*N* = 1849981). For a user following Right-leaning news media more than the other types, following Republican members more than Democrat members and following Christian religious accounts more than other religions have a highly negative associated *β* values. On the other hand, users following Hindu religious accounts more than other religions, following left-leaning and mixed news media more than other types and medical associations and institutes more than the individual doctors have a highly positive estimate value.

**Table 2.**
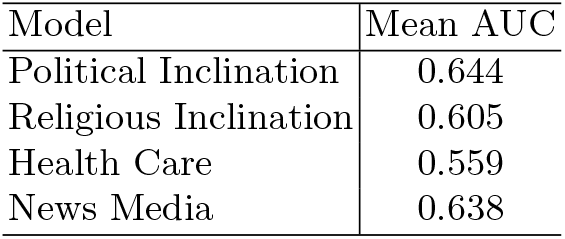
Mean Area Under Curve of logistic regression models for 20% held out test set and training set for 100 replicates.

| Model | Mean AUC |
| --- | --- |
| Political Inclination | 0.644 |
| Religious Inclination | 0.605 |
| Health Care | 0.559 |
| News Media | 0.638 |

## 4 Discussion and Related Work

As of June 2020, there have been numerous COVID-19 vaccine candidates undergoing trails [11]. Most estimates expect that a COVID-19 vaccine would be ready for distribution by the end of 2020. In such a scenario, the anti-vaxxers could pose a threat to a successful vaccination campaign. In this paper, we classified Twitter users talking about vaccines in the COVID-19 debate. We find that 25% of users show activity akin to that of anti-vaxxers. This is broadly consistent with the findings of a recently conducted survey by AP-NORC, which concluded that 20% of the surveyed population was not willing to get the future COVID-19 vaccine and 31% were not sure [25]. Furthermore, a study by Johnson and colleagues [16] on Facebook groups suggested that the anti-vaccination groups are attracting new users at a much higher rate compared to the pro-vaccination group. Although we primarily analyze COVID-19 messages tweeted by English speaking Twitter users, however, recent studies have suggested that the increase in the number of anti-vaxxers is not just limited to the English speaking countries [14].

To counter the growing number of anti-vaxxers, the scientific community needs to come up with appropriate messaging catering to the beliefs of the group. People tend to “persuade themselves to change attitude and behavior” [24] and hence communicators should tailor messaging based on the beliefs of audience groups [18]. Our findings suggest that bi-partisan messaging towards the effectiveness of vaccination is needed, more so for the followers of members of the US Congress from the southern and mid-western US. Members of the US Congress should provide consistent pro-vaccination messaging, such bi-partisan messaging has shown to reduce polarization [7]. Besides, we provide evidence that anti-vaxxers tend to follow more medical institutions than individual doctors. Hence, medical institutions should play a critical role in removing biases around scientific facts related to vaccination.

Previous studies have looked at the linguistic cues and bot activity in anti-vaxxers and vaxxers debate. A study by Memon et al. [21] concluded that anti-vaxxers and vaxxers use different linguistic cues related to intensifiers, pronouns, and uncertainty word. Broniatowski and colleagues [8] used fuzzy trace theory to prove that articles expressing bottom-line gist were more likely to be shared during the 2014-2015 Disneyland measles outbreak. In a different study, Broniatowski and colleagues [9] looked at the activity of bot-like accounts and Russian trolls in the vaccine debate. The study found that bot-like accounts and Russian trolls tweeted about vaccination at a higher rate. For appropriate messaging a need arises to look at the underlying constructs that could be nourishing and creating a bigger divide. In this paper, we focus on the difference in beliefs based on the followership of partisan, religious, medical communicators, and news media.

We explore the partisan divide between vaxxers and anti-vaxxers. A recent study about vaccine debate from the year 2015 to 2017 by Walter et al. suggested that the vaccine debate is increasingly getting polarized around political prejudice [27]. We further provide evidence of this finding. Moreover, we provide evidence of the partisan divide in the news sources followed by vaxxers and anti-vaxxers. Anti-vaxxers are more likely to follow right-leaning news media as compared to left-leaning or news media with mixed audiences. Hence, right-leaning news media should provide consistent and strong pro-vaccination messaging. Although the prediction power of the news media logistic model was similar to the political inclination model, we see a higher negative regression coefficient (*β*) value of right-leaning news media compared to Republican *β* value. Indicating that a user is more likely to be anti-vaxxer if the user follows right-leaning media more than other media types than compared to if the user follows Republican congress members more than Democrat congress members.

In this study, we provide evidence that Russian, Chinese, and Iranian state-owned news sources have a higher variation of audience beliefs compared to news sources owned by other countries. Some of the state media from these countries have a higher fraction of anti-vaxxers than compared to other state media from the same country. Further analysis is required to delineate the link between state media messaging and vaccine beliefs.

Furthermore, we provide evidence that some of the popular Christianity related accounts are followed more by anti-vaxxers than vaxxers. This could also be seen from the negative *β* values of Christianity associated variable. Interestingly, accounts related to Hinduism are followed more by vaxxers than anti-vaxxers.

Nevertheless, our analysis has several limitations. Our COVID-19 data stream is biased towards English speaking users. This could especially bias our results related to religious inclination and state media. Also, in our analysis for state media, religious accounts, and healthcare, we collected the most followed accounts in each group by Google and Twitter user search. However, these may not represent all the accounts in a particular group that are followed by vaxxers or anti-vaxxers.

## Data Availability

We will provide the list of accounts from each group used in our analysis on https://kilthub.cmu.edu/ as part of reviewed publication.

## Supplementary 1

### Model Evaluation

We evaluate our stance labels by taking a large sample and manually checking each account. We randomly sampled 500 users labeled as vaxxers and 500 users labeled as anti-vaxxer. Then we checked each user’s tweets in our dataset to assign a gold label. No Tweet in the randomly sampled users contained the seed hashtags used in §2.2. We label a user anti-vaxxer if the user is skeptical of vaccine science and vaxxer if we find no evidence that the user is skeptical. We report the confusion matrix in Table 3. The average precision of our stance detection model is 82.6%. The decrease in average precision is driven mainly by higher number of accounts that are predicted as anti-vaxxers but no evidence in their tweets suggested the same. Nevertheless, if the precision holds steady for all other users then the model accuracy is reasonable for our general conclusions.

**Table 3.** Confusion Matrix for the manual evaluation of stance detection algorithm.

|  | True Vaxxer | True Anti-Vaxxer |
| --- | --- | --- |
| Predicted Vaxxer | 441 | 59 |
| Predicted Anti-Vaxxer | 115 | 385 |

**Table 4.** The estimates (*β*) and standard error values of political inclination model as defined in §2.3.

|  | Estimates | Std. Errors |
| --- | --- | --- |
| Intercept | 1.5348 | 0.0066 |
| Democrat | -0.0267 | 0.0055 |
| Republican | -2.0430 | 0.0053 |
| Bot Score | 0.5834 | 0.0105 |
| $\log(\text{follows} + 1)$ | -0.0734 | 0.0011 |

**Table 5.** The estimates (*β*) and standard error values of religious inclination model as defined in §2.3.

|  | Estimates | Std. Errors |
| --- | --- | --- |
| Intercept | 1.5046 | 0.0065 |
| Hindu | 1.5479 | 0.0108 |
| Islam | 0.2281 | 0.0081 |
| Christian | -0.9892 | 0.0062 |
| Judaism | -0.1046 | 0.0095 |
| Bot Score | 0.4991 | 0.0101 |
| $\log(\text{follows} + 1)$ | -0.1130 | 0.0010 |

**Table 6.** The estimates (*β*) and standard error values of healthcare model as defined in §2.3

|  | Estimates | Std. Errors |
| --- | --- | --- |
| Intercept | 1.7305 | 0.0064 |
| Doctors | -0.0059 | 0.0095 |
| Medical Assc. & Inst. | 0.2072 | 0.0049 |
| Bot Score | 0.3496 | 0.0099 |
| $\log(\text{follows} + 1)$ | -0.1465 | 0.0010 |

**Table 7.** The estimates (*β*) and standard error values of news media based on the Pew Research survey [17] as defined in §2.3

|  | Estimates | Std. Errors |
| --- | --- | --- |
| Intercept | 1.6603 | 0.0067 |
| Left News | 0.1960 | 0.0047 |
| Right News | -2.9724 | 0.0085 |
| Mixed News | 0.1036 | 0.0121 |
| Bots Score | 0.4200 | 0.0106 |
| $\log(\text{follows} + 1)$ | -0.1024 | 0.0011 |

### Regression Model Estimates

## Footnotes

1 https://developer.Twitter.com/en/docs/tweets/search/overview/standard

2 We primarily use english and WHO terms - “coronavirus”, “coronaravirus”, “wuhan virus”, “wuhanvirus”, “2019nCoV”, “NCoV”, “NCoV2019”, “covid”

3 We use the parameter values as defined in [19] as {*k* = 5000, *p* = 5000, *θ*^*I*^ = 0.1, *θ*^*U*^ = 0.0, *θ*^*T*^ = 0.7}.

4 We will provide the list of accounts from each group used in our analysis on https://kilthub.cmu.edu/ as part of reviewed publication.

5 We collected followers of each parliamentarian from May 1st to May 15th, 2020. During this time period, we also collected followers of accounts related to religious inclination, healthcare, and news media.

6 Although we were able to collect close to 200 accounts for each religion, for our analysis we only use the top 150 most followed accounts from each religion.

7 Certain news sources could have multiple accounts serving a particular market, for example FoxSports, NytimesArts, etc. In our analysis, we focus on the main Twitter accounts whose name matches exactly with the ones reported in Pew Research’s report.

